# Comparable specimen collection from both ends of at-home mid-turbinate swabs

**DOI:** 10.1101/2020.12.05.20244632

**Authors:** Melissa Truong, Brian Pfau, Evan McDermot, Peter D. Han, Elisabeth Brandstetter, Matthew Richardson, Ashley E. Kim, Mark J. Rieder, Helen Y. Chu, Janet A. Englund, Deborah A. Nickerson, Jay Shendure, Christina M. Lockwood, Eric Q. Konnick, Lea M. Starita, on behalf of Seattle Flu Study investigators

## Abstract

Unsupervised upper respiratory specimen collection is a key factor in the ability to massively scale SARS-CoV-2 testing. But there is concern that unsupervised specimen collection may produce inferior samples. Across two studies that included unsupervised at-home mid-turbinate specimen collection, ∼1% of participants used the wrong end of the swab. We found that molecular detection of respiratory pathogens and a human biomarker were comparable between specimens collected from the handle of the swab and those collected correctly. Older participants were more likely to use the swab backwards. Our results suggest that errors made during home-collection of nasal specimens do not preclude molecular detection of pathogens and specialized swabs may be an unnecessary luxury during a pandemic.

---

At-home respiratory specimen collection for pathogen testing enables community sampling. Furthermore, it requires neither a healthcare worker’s time nor personal protective equipment and symptomatic individuals can continue to self-isolate. However, questions remain as to whether unsupervised upper respiratory specimen collection by individuals in their homes reliably produce specimens that are of high enough quality for pathogen testing. From October 2019 through May 2020, the Seattle Flu Study (1, 2) and the greater Seattle Coronavirus Assessment Network (SCAN, scanpublichealth.org) screened 16,785 mid-turbinate swabs that were self-collected by participants at home for respiratory pathogens. The at-home kits contained a flocked, mid-turbinate swab (Copan 56380CS01 or 56750CS01) - either adult or pediatric, a tube of universal transport media (UTM), and instructions on how to self-collect a specimen or collect a specimen for a child and return it to the lab (2). Of the kits distributed to individuals in the Seattle metropolitan area, most resulted in swabs returned appropriately according to the instructions in the kit, but 138/16,785 (0.8%) kits were returned to the lab with the swab handle in the UTM tube rather than the swab itself. The swab handle is non-tapered, hard plastic with decreased surface area compared to the flocked end of the swab **(Fig 1A)**. We were puzzled by this phenomenon, and sought to evaluate whether handle-collected specimens were comparable to flocked swabs themselves for molecular pathogen detection. We also assessed demographic covariates associated with errors in swab collection.

**Figure 1.**
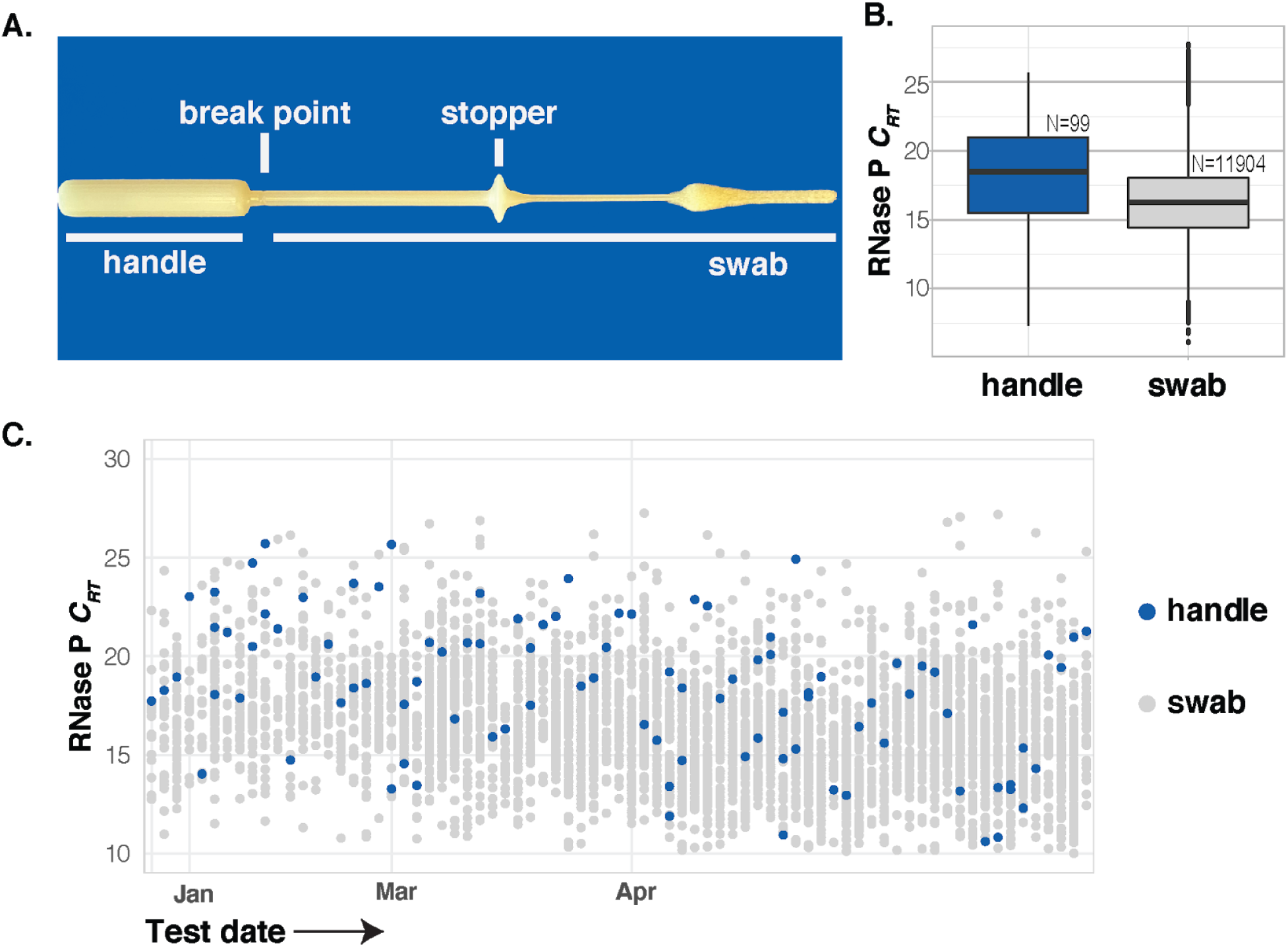
RNase P levels from either end of mid-turbinate swabs. **A**. Mid-turbinate swab (Copan 56380CS01), underlined handle or swab was placed in UTM by participants. **B**. *C*_*RT*_ values from all samples with RNase P detected, dashed line indicates detection limit. **C**. *C*_*RT*_ values for human RNAse P among batches of specimens (arranged on the x-axis by date) where at least one handle specimen was used.

Of the 16,782 specimens, 12,006 were analyzed for the presence of 24 respiratory pathogens using our Taqman-based detection panel including the 99 specimens collected with the handle (**Table 1 and supplementary material**). Samples collected after January 1, 2020 were additionally tested for the presence of SARS-CoV-2 using a separate RT-PCR assay. As a quality control metric to determine if a sufficient nasal specimen was collected for each sample, both assay platforms measure the amount of human RNase P. Specimens with RNase P relative cycle threshold (*C*_*RT*_) > 28 were considered to be a failed collection. The failure rate for all properly-collected specimens was 2.0% (238/12142). We expected a high failure rate for the handle collected specimens, however only 2.9% (3/102) failed this quality control metric, a non-significant difference (p = 0.46, Fisher’s exact test). The *C*_*RT*_ values for human marker RNase P for handle-collected specimens were higher than properly collected specimens (**Fig 1B**), with a mean *C*_*RT*_ value of 16.32 (95% CI 16.27 - 16.37) for swabs and 18.19 (95% CI 17.43 - 18.96) for handles (p < 0.01). However, the *C*_*RT*_ from handle-collected specimens generally fell within the same range and well below the failure threshold (**Fig 1C**), showing that the handles were indeed collecting human cells. In addition, we identified multiple respiratory pathogens, including SARS-CoV-2, at similar rates of detection with both swabs and swab handles (p=0.52) (**Table 1**).

**Table 1.**
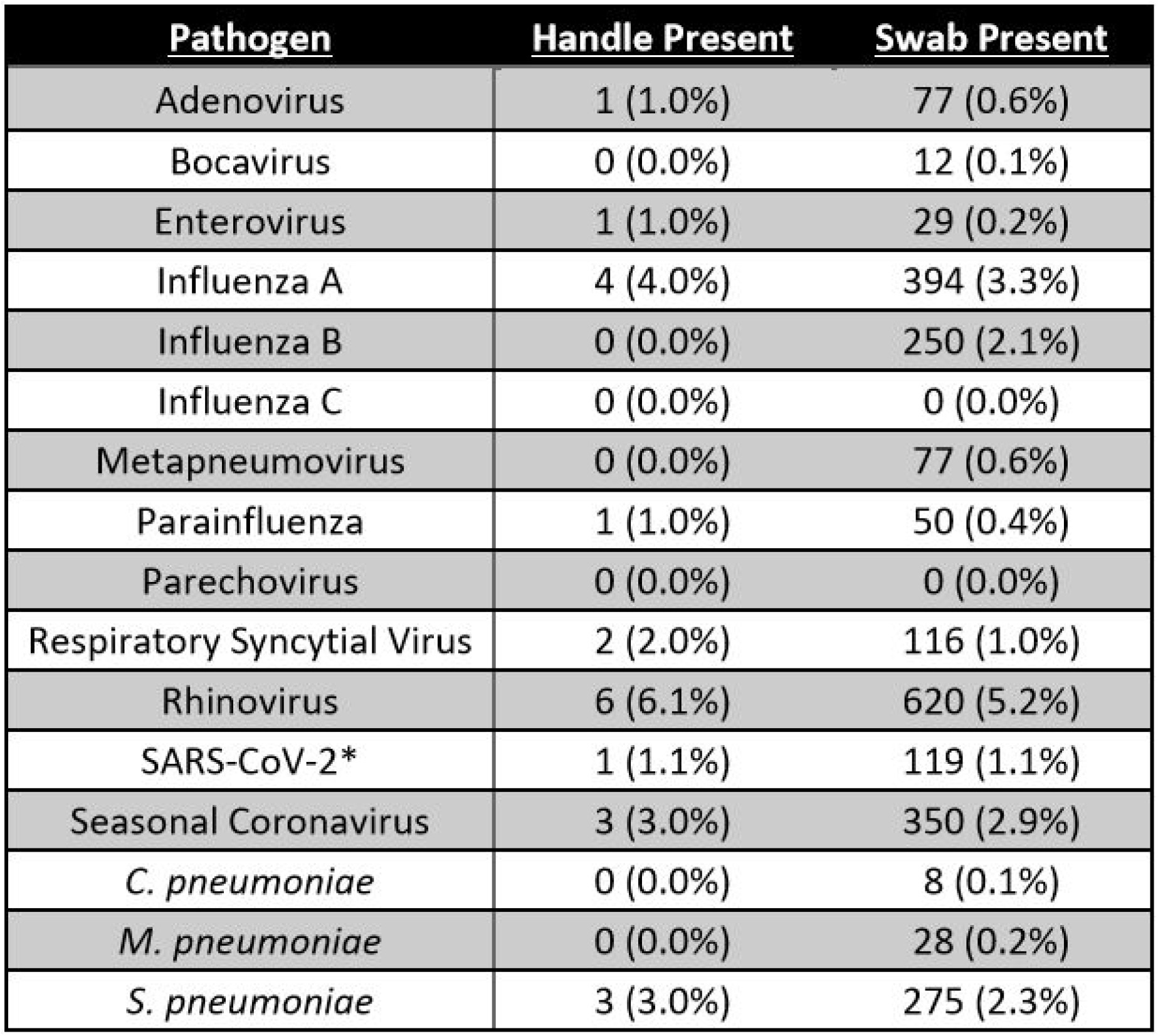
Detection rates of respiratory pathogens.

We examined the clinical data associated with the samples to determine which participants were more likely to collect a specimen with the handle. Participants who swabbed with the handle were more likely to be older (**Fig 2A**), with a median age of 62 compared to 39 for those who followed the instructions (p <0.01). There was no significant difference in handle use between men and women (p=0.22) or across income brackets (supplementary material). Interestingly, participants who had erroneously used the handle were more confident that they had collected a quality specimen (**Fig 2B**, 73% highly confident with the handle vs. 62% with the swab, p=0.02), and reported lower overall discomfort (**Fig 2C**, 42% reported no discomfort with the handle vs. 16% with the swab, p<0.01)

**Figure 2.**
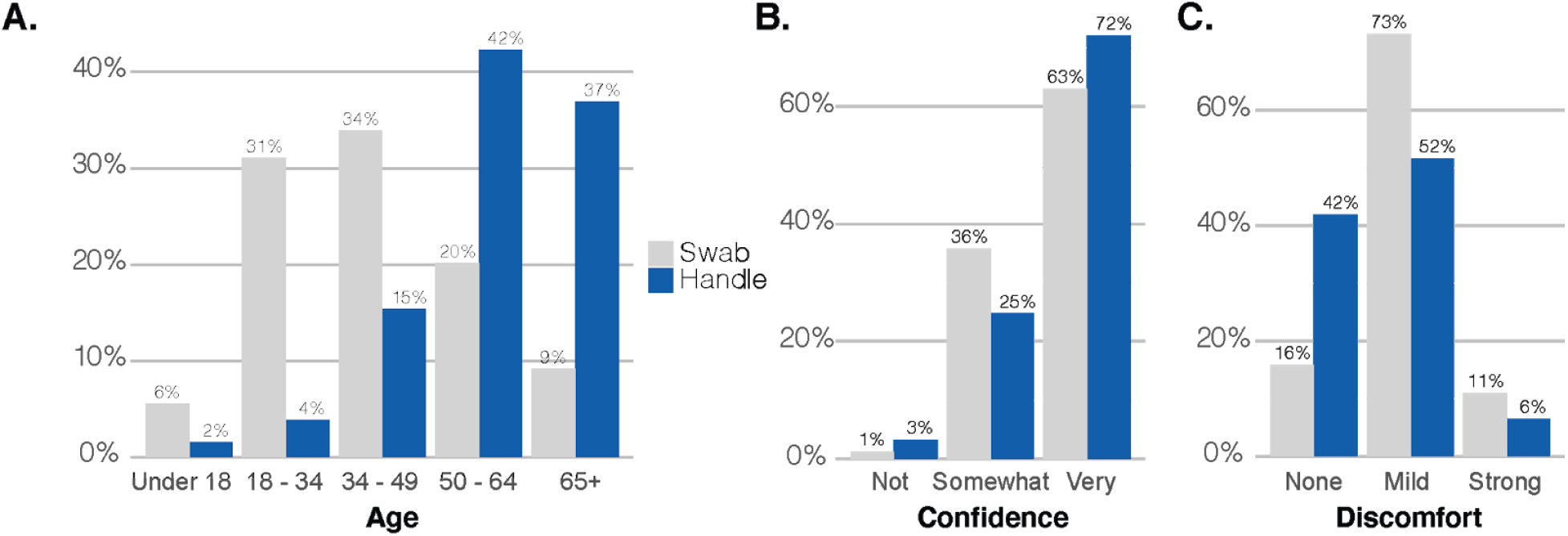
Specimen collection error by age and self reported confidence and discomfort. **A**. Age of participants, **B**. Self reported confidence in specimen collection and **C**. Self-reported discomfort during specimen collection by which end of the swab was used.

We investigated unanticipated operator error in two large studies employing at-home mid-turbinate swab collection, and determined that participants who used the plastic handle rather than flocked swab to collect their sample and submit it to a laboratory were able to collect an adequate nasal specimen for molecular detection of respiratory pathogens. Like other studies(3), these results suggest that the use of specialty swabs may only result in marginal increases in pathogen detection. They also suggest that even if participants do not closely adhere to instructions, they can still collect a sample that is sufficient for the molecular detection of respiratory pathogens including influenza and SARS-CoV-2. In this time of global swab shortages and an unabating pandemic, the type of swab used may not be critical for detection of SARS-CoV-2.

## Data Availability

All data is available, as is the code for data analysis and figure regeneration

## Acknowledgements

We would like to thank the Seattle Flu Study and SCAN participants for their invaluable contributions to this research, the entire Seattle Flu Study team for making this study possible and Katrina Van Raay for R code.

The Seattle Flu Study and SCAN are administered by the Brotman Baty Institute for Precision Medicine and funded by Gates Ventures, the private office of Bill Gates. The funder was not involved in the design of the study and does not have any ownership over the management and conduct of the study, the data, or the rights to publish. LMS and JS are funded by 1RM1HG010461-01 from the NHGRI and JS is an Investigator of the Howard Hughes Medical Institute. REDCap at ITHS is supported by the National Center for Advancing Translational Sciences of the National Institutes of Health under Award Number UL1 TR002319.

## Ethics Approval

The Seattle Flu Study received approval by the University of Washington’s Institutional Review Board at the (UW IRB; STUDY00006181) and informed consent was obtained prior to study enrollment. Participants participated in SCAN as part of public health surveillance.

## Seattle Flu Study Investigators Principal Investigators

Helen Y. Chu^1,7^, Michael Boeckh^1,2,7^, Jeffrey S. Duchin^10^, Janet A. Englund^3,7^, Michael Famulare^4^, Barry R. Lutz^5,7^, Deborah A. Nickerson^6,7^, Mark J. Rieder^7^, Lea M. Starita^6,7^, Matthew Thompson^9^, Jay Shendure^6,7,8^, and Trevor Bedford^2,6,7^

## Affiliations

1. Department of Medicine, University of Washington
2. Vaccine and Infectious Disease Division, Fred Hutchinson Cancer Research Center
3. Seattle Children’s Research Institute
4. Institute for Disease Modeling
5. Department of Bioengineering, University of Washington
6. Department of Genome Sciences, University of Washington
7. Brotman Baty Institute For Precision Medicine
8. Howard Hughes Medical Institute
9. Department of Family Medicine, University of Washington
10. Public Health – Seattle King County

## Appendix

### Study description

The Seattle Flu Study Swab & Send program was IRB approved (STUDY00006181) and consent is obtained from participants or their guardians online at time of enrollment.The greater Seattle Coronavirus Assessment Network (SCAN) is a public health initiative under the direction of Public Health Seattle King County and permission is obtained from participants for molecular testing. For both studies, participants were sent a self-test kit to their home following online enrollment and answering a brief questionnaire. Participants collected their own mid-turbinate specimen, unsupervised, with written and video instructions (scanpublicheatlh.org). Instructions and materials were included for the participants to package and ship the sample according to IATA bio-specimen regulations. An in-depth description of the Seattle Flu Study Swab & Send program can be found (2). SCAN is based on the Swab & Send program with improvements aimed at achieving greater geographic and demographic diversity.

### Molecular testing

Specimens were shipped to the Brotman Baty Institute for Precision Medicine via commercial couriers or the US Postal Service at ambient temperatures and opened in a class II biological safety cabinet in a biosafety level-2 laboratory. When opening specimen packages and transferring samples, technicians recorded basic information, including participant compliance with labelling and packaging instructions. Sample entries with notes indicating that mistakes had been made by participants were manually curated to determine if they belonged in the handle or the standard swab cohort. Samples with both the swab and the handle in the UTM vial were excluded from both groups.

Two or three 650 µL aliquots of UTM were collected from each specimen and stored at 4°C until the time of nucleic acid extraction, performed with the MagnaPure 96 small volume total nucleic acids kit (Roche).

Molecular assays were performed at the Northwest Genomics Center (Department of Genome Sciences, the University of Washington). Extracted nucleic acids were tested for the presence of 24 respiratory pathogens by TaqMan RT-PCR on the OpenArray platform (**Appendix Table 1**) and separate RT-PCR assay for SARS-CoV-2. For the Open Array pathogen panel, the extracted nucleic acid samples were added to a PreAmp reaction master mix containing TaqPath 1-Step RT-qPCR Master Mix CG, a custom TaqMan PreAmp oligonucleotide pool, and a spike-in control (TaqMan Universal Xeno RT control, ThermoFisher). The pre-amplification reactions were reverse transcribed and amplified for 14 PCR cycles according to the manufacturer’s recommendations. The preamplified samples were diluted and added to Open Array Mastermix (ThermoFisher); the mix was then re-arrayed onto custom Open Array plates containing each RT-PCR assay in duplicate. RT-PCR was performed on a QuantStudio 12 (Applied Biosystems), with cycling parameters set according to the manufacturer’s recommendations, for a total of 35 cycles. Positive and negative template controls were included in each extraction and PCR batch.

**Table 1.**
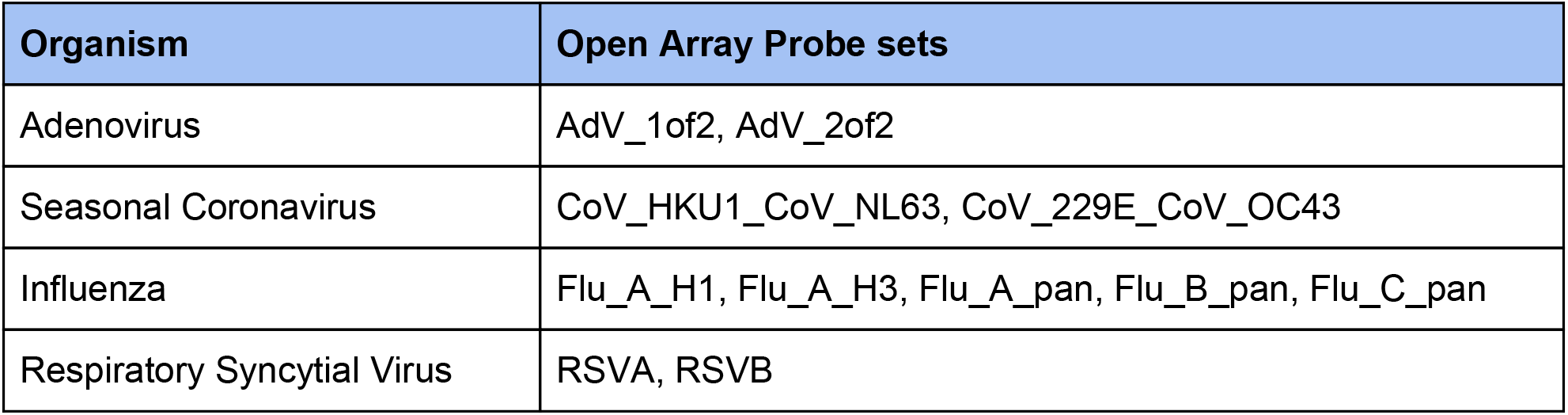
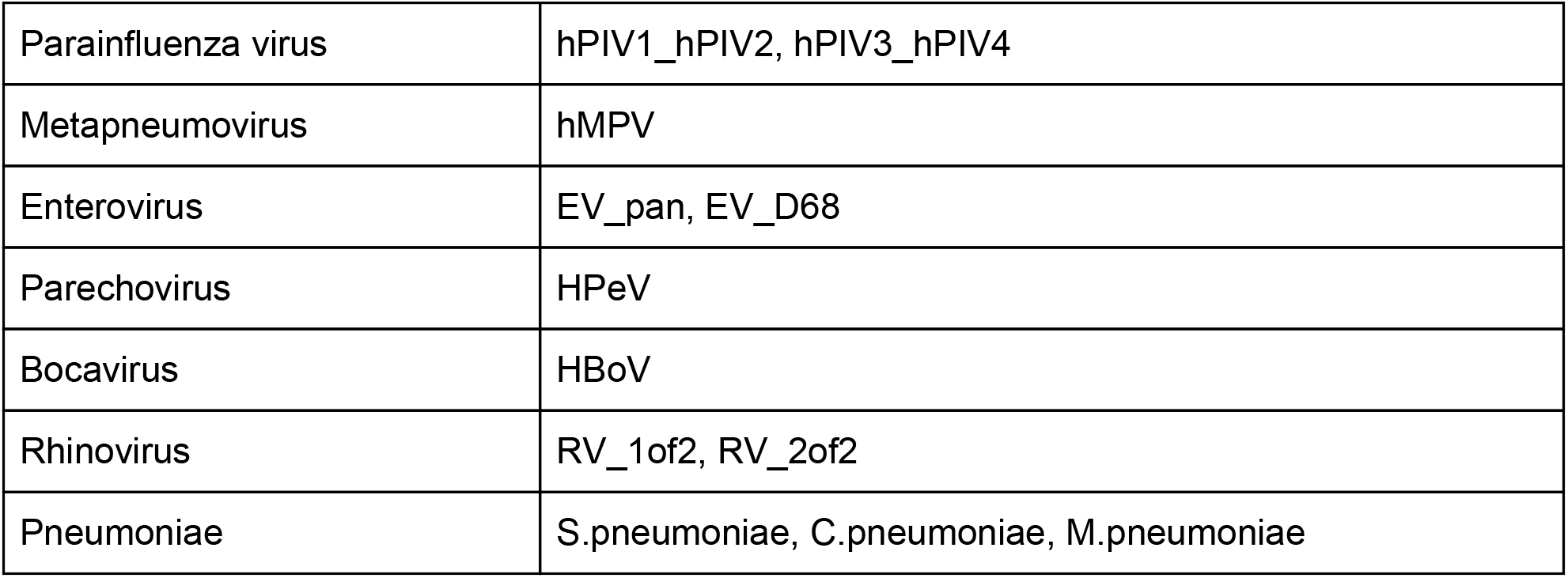

Data files from the Open Array were imported into and processed through our in-house LIMS system. Detection of each technical replicate is calculated independently using threshold values provided by ThermoFisher; the relative cycle threshold (*C*_*RT*_) must be less than or equal to the *C*_*RT*_ threshold for each respiratory pathogen to be deemed present. Additionally, both the C_Q_ Conf and Amp Score must be greater than or equal to separate thresholds for a respiratory pathogen to be flagged as detected. Samples with an RNAse P *C*_*RT*_ > 28 are considered failed. The failure rate for all properly-collected specimens versus handle-collected specimens was analyzed via R Studio (Version 1.2.5033) using Fisher’s exact test.

SARS-CoV-2 was detected using a laboratory-developed test (LDT) or research assay. For the LDT, SARS-CoV-2 detection was performed using real-time RT-PCR with a probe sets targeting Orf1b and S with FAM fluor (Life Technologies 4332079 assays # APGZJKF and APXGVC4APX) multiplexed with an RNaseP probe set with VIC or HEX fluor (Life Technologies A30064 or IDT custom) each in duplicate on a QuantStudio 6 instrument (Applied Biosystems). The research assay employs only the Orf1b and RNaseP multiplexed RT-PCR in duplicate. Three or four replicates for RNase P and SARS-CoV-2 were required to have a cycle threshold (*C*_*T*_) < 40 for a sample to be considered positive in the LDT or both replicates must be positive in the research assay. Specimens with two replicates with SARS-CoV-2 detected are considered inconclusive.

### Data Analysis

RNAse P *C*_*T*_ values and participant demographics were analyzed in RStudio Version 1.2.5042. Replicate *C*_*T*_ values for RNAse P were averaged, and the mean *C*_*T*_ values for handles and swabs were compared using a two-tailed Welch’s two-sample t-test. Pathogen detection rates were compared using a paired t-test.

Participant data were collected and managed using REDCap electronic data capture tools hosted at the University of Washington (4, 5). REDCap (Research Electronic Data Capture) is a secure, web-based software platform designed to support data capture for research studies, providing 1) an intuitive interface for validated data capture; 2) audit trails for tracking data manipulation and export procedures; 3) automated export procedures for seamless data downloads to common statistical packages; and 4) procedures for data integration and interoperability with external sources. Participants measured their level of discomfort and confidence in their swab technique on a voluntary online survey taken at time of sample collection. The three levels of comfort and confidence, respectively, were each compared using a Fisher’s exact test. Participant age was compared using a two-tailed Welch’s two-sample t-test, and sex was compared with a Pearson’s Chi-squared test with Yates’ continuity correction.

