## Supplementary File 2 for "Comparable specimen collection from both ends of at-home mid-turbinate swabs"

#### SECTION 0: LOADING YOUR LIBRARY AND DATASET

Load the required packages below:

```
library(tidyverse)
library(data.table)
library(scales)
```

Remember to set your working directory to the appropriate path.

A CSV file is loaded and read:

1. This file includes the raw dataset of all SCAN and SFS samples tested for SARS-CoV-2.

Once loaded, a dataframe called **SCAN_SFS_df** is created. This dataframe will be used for all of our analysis.

Our four analyses are:

1) Comparison of Failure Rate
2) Comparison of Participant Data
3) Comparison of RNAse P Ct Values
4) Comparison of Pathogen Detection

Additionally, we obtain numbers of total samples, total handle-type samples, total samples tested for each pathogen, and total samples tested for RNAse P.

```
# dataframe for the raw SCAN/SFS dataset
SCAN_SFS_df = tbl_df(read.csv("supplementary_file_1.csv"))
# total number of samples screened
screened_swabs = nrow(SCAN_SFS_df)
# total number of handle-type
handle_type = filter(SCAN_SFS_df, side_used == "handle")
handle_type_count = nrow(handle_type)
# total samples tested for pathogen detection
path_detect = filter(SCAN_SFS_df, !is.na(detected))
path_detect_count = nrow(path_detect)
# total samples tested for RNAse P detection
rnasep_detect = filter(SCAN_SFS_df, !is.na(rnasep_detected))
rnasep_detect_count = nrow(rnasep_detect)
```

There are:

1. **16,782 total samples screened**
2. **138 handle-type samples**
3. **12,006 samples tested for pathogens on our Taqman-based detection panel**
4. **12,244 samples tested for RNAse P**

### SECTION 1: COMPARISON OF FAILURE RATES

#### Creating the Failure Rate Dataframe

From the SCAN_SFS_df dataframe, we will create a new dplyr dataframe specifically for failure rate calculation.

```
failure_rate_df <- SCAN_SFS_df

# selecting the needed columns
failure_rate_df <- select(SCAN_SFS_df, sample_id, side_used, rnasep_detected)

# filter out NA target status
failure_rate_df  <- filter(failure_rate_df, !is.na(rnasep_detected)
```

Failure rates can be compared using this dataframe.

#### Creating The Failure Rate Contingency Table

A contingency table is calculated to compare the number of detected versus not detected samples for each side used, handle or swab.

```
# create table based on type - handle/swab and target status - detected/not detected
handle_swab_table <- table(failure_rate_df$side_used, failure_rate_df$rnasep_detected)

# add sum of handle and swab sample to table
handle_swab_table_w_sum <- addmargins(table(failure_rate_df$side_used,
                                            failure_rate_df$rnasep_detected), 2)

# view table on console
handle_swab_table
```

The table with the sum should appear as:

| | Detected | Not Detected | Sum |
|---|---|---|---|
| handle | 99 | 3 | 102 |
| swab | 11904 | 238 | 12142 |

#### Calculating Significant Difference between Failure Rates

A Fisher's exact test is used to determine whether the failure rates differ by handle versus swab.

```
failure_rate_p_value <- fisher.test(handle_swab_table)
failure_rate_p_value
```

With a p-value of **0.46**, we do not find a significant difference between the two failure rates.

### SECTION 2: ANALYZING PARTICIPANTS' DATA

#### Creating the Participants' Data Dataframe

From the SCAN_SFS_df dataframe, we will create a new dplyr dataframe specifically for participant data analysis.

```
participant_df <- SCAN_SFS_df
```

#### Performing Fisher's Test for Confidence and Discomfort During Nasal Swab Collection

Participant confidence that the sample was taken correctly and discomfort while taking the sample are compared using a Fisher's exact test.

```
# create a tables of confidence
confidence_table <- table(participant_df$confidence, participant_df$side_used)
confidence_prop <- prop.table(table(participant_df$confidence, participant_df$side_used))

# create a tables of discomfort
discomfort_table <- table(participant_df$discomfort, participant_df$side_used)
discomfort_prop <- prop.table(table(participant_df$discomfort, participant_df$side_used))

# calculate p-value for confidence of handle versus swab
fisher.test(participant_df$confidence, participant_df$side_used)

# calculate p-value for discomfort for handle versus swab
fisher.test(participant_df$discomfort, participant_df$side_used)

# binding data of confidence and discomfort together
confidence_prop_table <- data.table(confidence_prop)
colnames(confidence_prop_table) <- c("confidence","side_used","freq")
discomfort_prop_table <- data.table(discomfort_prop)
colnames(discomfort_prop_table) <- c("discomfort","side_used","freq")
nasal_swab_usability_table <- bind_rows(confidence_prop_table,discomfort_prop_table)

# final table is saved as a csv
write_csv(nasal_swab_usability_table,"nasal_swab_usability_table.csv")
```

The p-value for confidence between handle versus swab is **0.01**.

The p-value for discomfort between handle verus swab is $<$ **0.01**.

nasal__swab__usability__table is saved as a csv into the path directory. This table, categorized between handle-type versus swab-type, includes the frequency of participants' answers towards confidence and discomfort.

#### Creating Bar Plots of Confidence and Discomfort

To visualize the data using a bar plot, relative frequency must be calculated for either side used. Then, a bar plot showing the percentage of participants' degree of confidence is generated.

```
# create table with relative frequency
rel_confidence_prop_table <- confidence_prop_table %>% group_by(side_used) %>%
  mutate(relfreq = freq / sum(freq))

# rename column names in prep of bar plot generation
library(plyr)
rel_confidence_prop_table$confidence <- revalue(rel_confidence_prop_table$confidence,
                                                c("not_con" = "Not \n confident",
                                                  "some_con" = "Somewhat \n confident",
                                                  "v_con" = "Very \n confident"))
detach("package:plyr", unload = TRUE)
rel_confidence_prop_table$side_used <- factor(rel_confidence_prop_table$side_used,
                                              levels = c("swab","handle"))

# create barplot for discomfort
ggplot(data = rel_confidence_prop_table,aes(x = confidence,y = relfreq,fill = side_used)) +
  geom_bar(stat="identity", position = "dodge") +
  scale_y_continuous(labels = scales::percent) +
  theme(panel.background = element_rect(fill = "white"),
        panel.grid.major.y = element_line(size = 0.5, linetype = 'solid', color = "grey"),
        panel.grid.major.x = element_blank(),
        legend.position = "none",
        plot.title = element_text(face = "bold", hjust = 0.5),
        axis.text.x = element_text(size = 12),
        axis.title.x = element_text(face = "bold",vjust= -1),
        axis.ticks = element_blank(),
        axis.title.y = element_blank())+
  geom_text(aes(label=scales::percent(relfreq,accuracy =1), y = relfreq),
            vjust=-0.5, size=3.5,position = position_dodge(width = 1)) +
  ggtitle("Participant Confidence that Sample was \n Taken Correctly by Handle and Swab") +
  scale_fill_manual(values = c("swab" = "lightgrey",
                               "handle" = "#095DA8")) +
  labs(x="Confidence", fill = "Side Used")

# removing unneeded dataframe
rm(confidence_prop_table)
```

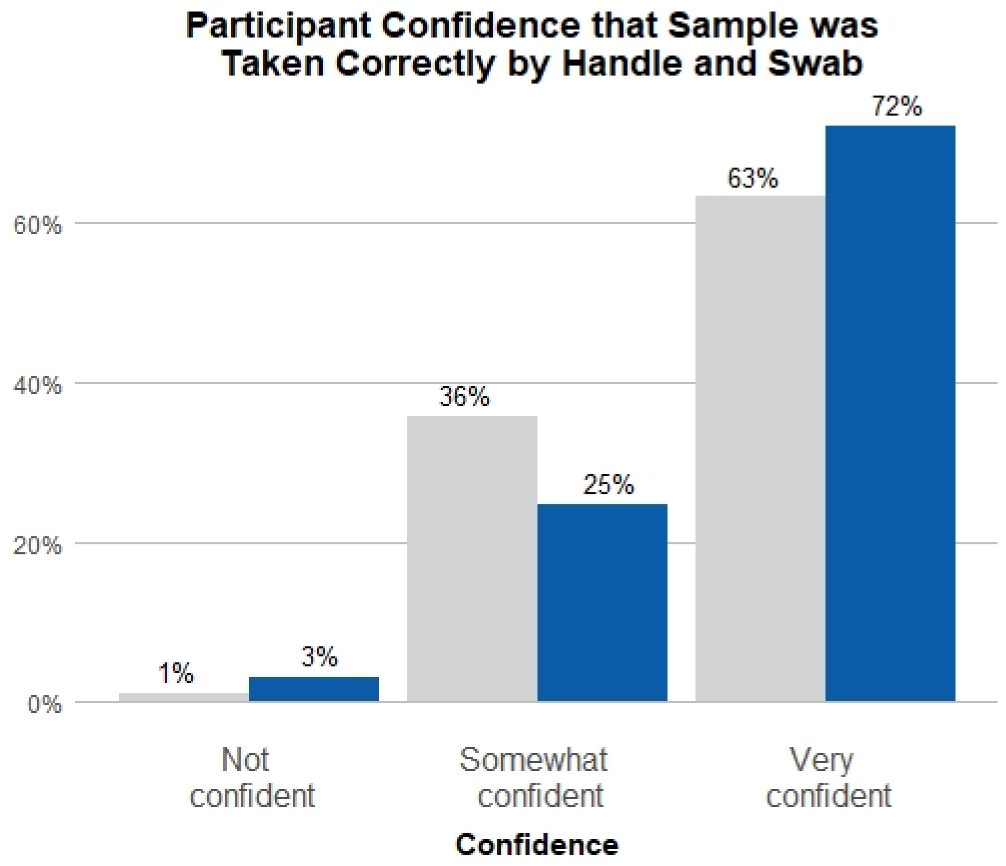


Similarly, a bar plot can be generated to visualize rate of discomfort between handle versus swab. Once again, a relative frequency column must be created.

```
# create table with relative frequency
rel_discomfort_prop_table <- discomfort_prop_table %>% group_by(side_used) %>%
  mutate(relfreq = freq / sum(freq))

# rename column names in prep of bar plot generation
library(plyr)
rel_discomfort_prop_table$discomfort <- revalue(rel_discomfort_prop_table$discomfort,
                                                c("no_dis" = "No \n discomfort",
                                                  "mild_dis" = "Mild \n discomfort",
                                                  "strong_dis" = "Strong \n discomfort"))
detach("package:plyr", unload = TRUE)
rel_discomfort_prop_table$discomfort <- factor(rel_discomfort_prop_table$discomfort,
                                               levels = c("No \n discomfort",
                                                          "Mild \n discomfort",
                                                          "Strong \n discomfort"))
rel_discomfort_prop_table$side_used <- factor (rel_discomfort_prop_table$side_used,
                                               levels = c("swab","handle"))

# create barplot for discomfort
ggplot(data = rel_discomfort_prop_table,aes(x = discomfort, y = relfreq, fill = side_used))+
  geom_bar(stat="identity", position = "dodge")+
  scale_y_continuous(labels = scales::percent)+
```

```r
  theme(panel.background = element_rect(fill = "white"),
        panel.grid.major.y = element_line(size = 0.5, linetype = 'solid', color = "grey"),
        panel.grid.major.x = element_blank(),
        legend.position = "none",
        plot.title = element_text(face = "bold", hjust = 0.5),
        axis.text.x = element_text(size = 12),
        axis.title.x = element_text(face = "bold",vjust= -1),
        axis.ticks = element_blank(),
        axis.title.y = element_blank())+
  geom_text(aes(label=scales::percent(relfreq, accuracy = 1), y = relfreq),
            vjust = -0.5, size=3.5, position = position_dodge(width = 1)) +
  ggtitle("Discomfort Experienced by Participants by \n Handle and Swab") +
  scale_fill_manual(values = c("swab" = "lightgrey",
                               "handle" = "#095DA8")) +
  labs(x="Discomfort", fill = "Side Used")

# removing unneeded dataframe
rm(discomfort_prop_table)
```

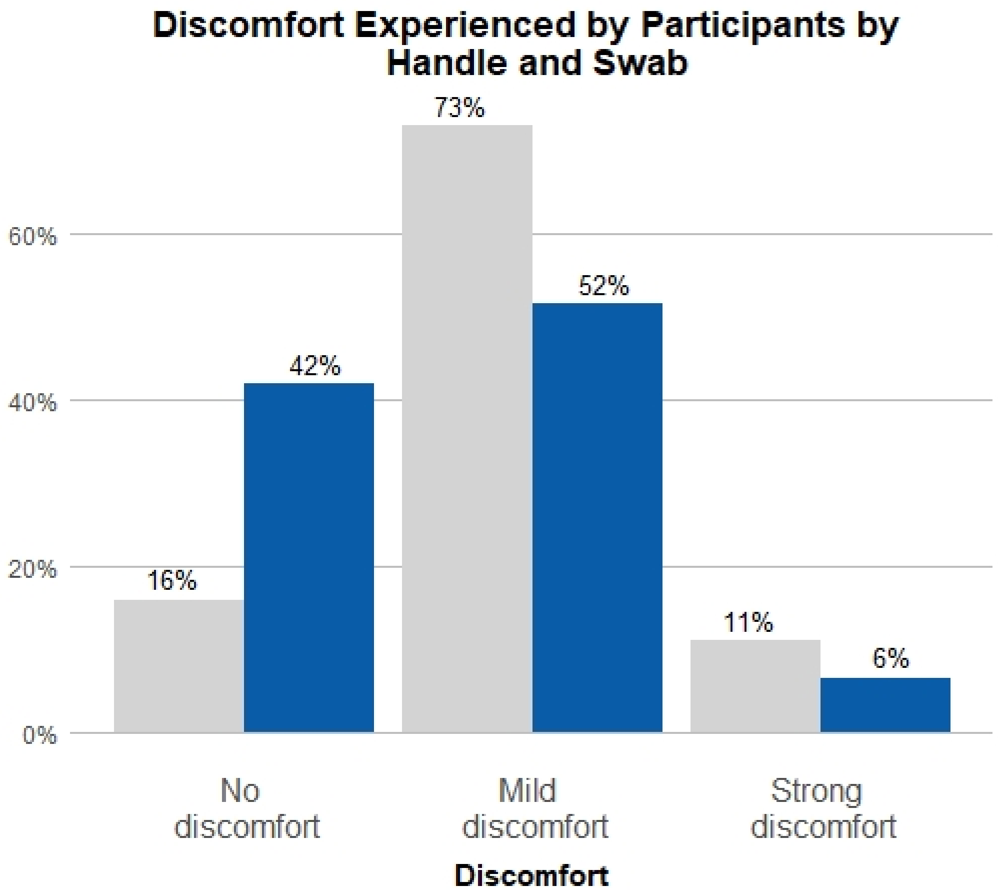


#### Data Analysis and Visualization for Age

The mean and median age between handle and swab can be evaluated by grouping by side used and then summarizing the mean and median for each group.

```
participant_df %>%
  dplyr::group_by(side_used) %>%
  dplyr::summarise(mean = mean(age,na.rm=TRUE),median = median(age,na.rm=TRUE), n = n())
```

The mean and median age for handle is **58.7** and **62** respectively. The mean and median age for swab is **41.2** and **39** respectively.

Normality is assumed for a dataset of this size, so a two-sample t-test can be used for the difference in mean age between handle and swab.

```
# t test for difference in age
dfAge <- data.frame(side_used = participant_df$side_used, age = participant_df$age)
dfAge <- dfAge %>% mutate(id = row_number())
age_t_test <- dfAge %>% spread(side_used,age)
t.test(age_t_test$handle,age_t_test$swab)
```

The p-value for the t-test $< \mathbf{0.01}$.

Age difference between handle and swab is visualized using a histogram. First, age is separated into the appropriate binning.

```
# all the data on age is separated into age binnings and NAs are omitted
agedata <- participant_df %>% filter (!is.na(age))
age_group <- agedata %>% mutate(age_group = case_when(
  age >= 65 ~ '65+',
  age >= 50 ~ '50-64',
  age >= 35 ~ '35-49',
  age >= 18 ~ '18-34',
  age < 18 ~ 'Under 18'))

# create new dataframe with age binnings
age_group2 <- data.frame(age = age_group$age_group, side_used = age_group$side_used)

# create new dataframe with age binnings and frequency
age_hist <- age_group2 %>%
  group_by(side_used,age,.drop=FALSE) %>%
  dplyr::summarise(n = n()) %>%
  complete(side_used, fill=list(count_a =0))%>%
  mutate(freq = n / sum(n))%>%
  ungroup()

# QC-ing of age_hist, including adding a percentage column
age_hist$age <- factor(age_hist$age, levels = c("Under 18","18-34","35-49","50-64", "65+"))
age_hist$side_used <- factor (age_hist$side_used, levels = c("swab","handle"))
age_hist$percent <- scales::percent(age_hist$freq)
```

A histogram for participants' age can now be generated.

```
# histogram is generated
ggplot(age_hist, aes(x=age,y=freq,fill=side_used))+
  geom_bar(stat="identity",position = position_dodge(preserve="single"))+
  scale_y_continuous(labels = c("0%","10%","20%","30%","40%"),
                     breaks = c(0,0.1,0.2,0.3,0.4)) +
```

```r
theme(panel.background = element_rect(fill = "white"),
      panel.grid.major.y = element_line(size = 0.5, linetype = 'solid', color = "grey"),
      panel.grid.major.x = element_blank(),
      legend.position = "right",
      plot.title = element_text(face = "bold", hjust = 0.5),
      axis.text.x = element_text(size = 12),
      axis.title.x = element_text(face = "bold",vjust = -1),
      axis.ticks = element_blank(),
      axis.title.y = element_blank()) +
geom_text(aes(label=scales::percent(freq,accuracy = 1),y = freq), vjust = -0.5,
          size=3.5, position = position_dodge(width = 1)) +
ggtitle("Age of Participants by Handle and Swab") +
scale_fill_manual(values = c("swab" = "lightgrey",
                             "handle" = "#095DA8")) +
labs(x="Age", fill = "Side Used")
```

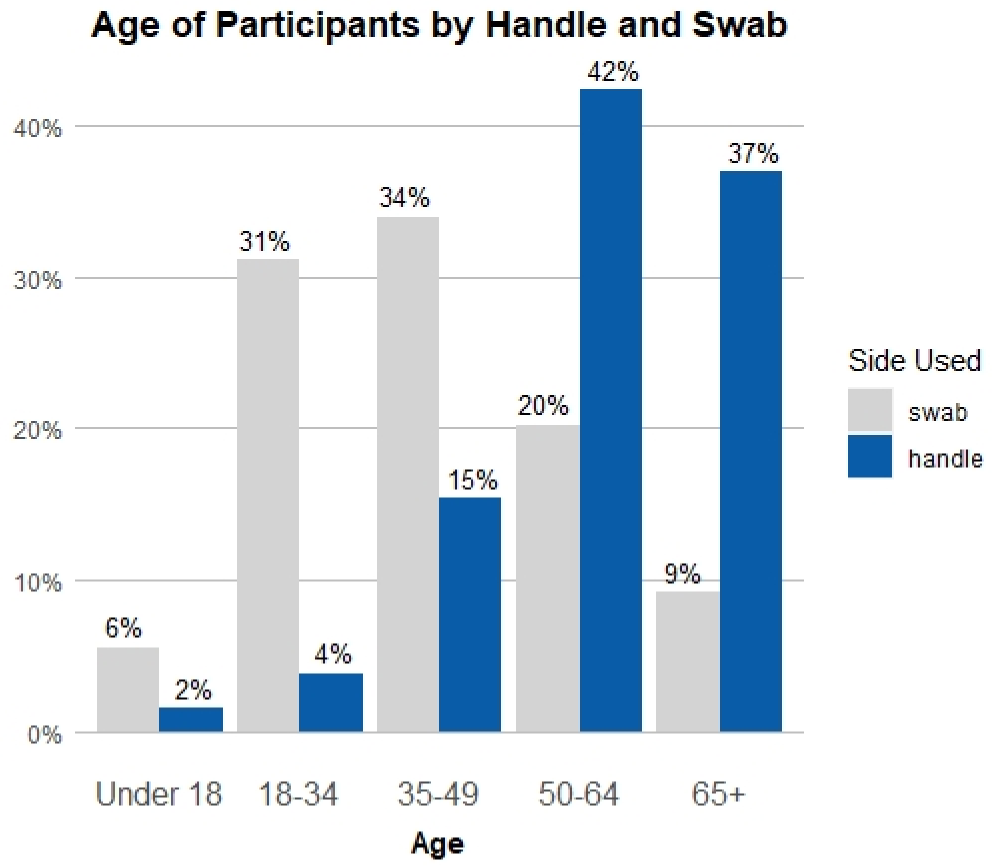


#### Data Analysis and Visualization for Sex

Before a bar plot can be generated, participants' sex are labeled and samples without sex data are removed. For the purpose of this analysis, participants with a non-binary gender or who did not report sex are excluded.

```r
# sex binning is created
sex_group <- participant_df %>% mutate(sex_group = case_when(
```

```
  sex == "male" ~ 'Male',
  sex == "female" ~ 'Female',
  sex == "other" ~ 'NA',
  sex == "dont_say" ~ 'NA'))

# NAs are removed
sex_group <- sex_group[!is.na(sex_group$sex_group), ]
sex_group <- sex_group %>% filter(!sex_group %in% "NA")

# dataframe created and modified
sex_hist <- sex_group %>%
  group_by(side_used,sex_group,.drop=FALSE) %>%
  dplyr::summarise(n = n()) %>%
  complete(side_used, fill=list(count_a =0))%>%
  mutate(freq = n / sum(n))%>%
  ungroup()
sex_hist$side_used <- factor(sex_hist$side_used, levels = c("swab","handle"))
```

With this dataframe created, a bar plot can be now generated.

```
# a bar plot can be generated
ggplot(sex_hist, aes(x=sex_group,y=freq,fill=side_used))+
  geom_bar(stat="identity",position = position_dodge(preserve="single"))+
  scale_y_continuous(labels = scales::percent)+
  theme(panel.background = element_rect(fill = "white"),
        panel.grid.major.y = element_line(size = 0.5, linetype = 'solid', color = "grey"),
        panel.grid.major.x = element_blank(),
        legend.position = "right",
        plot.title = element_text(face = "bold", hjust = 0.5),
        axis.text.x = element_text(size = 12),
        axis.title.x = element_text(face = "bold",vjust= -1),
        axis.ticks = element_blank(),
        axis.title.y = element_blank())+
  geom_text(aes(label=scales::percent(freq,accuracy =1),y=freq), vjust=-0.5, size=3.5,position = positi
  ggtitle("Sex of Participants by \n Handle and Swab")+
  scale_fill_manual(values = c("swab" = "lightgrey",
                               "handle" = "#095DA8"))+
  labs(x="Sex", fill = "Side Used")
```

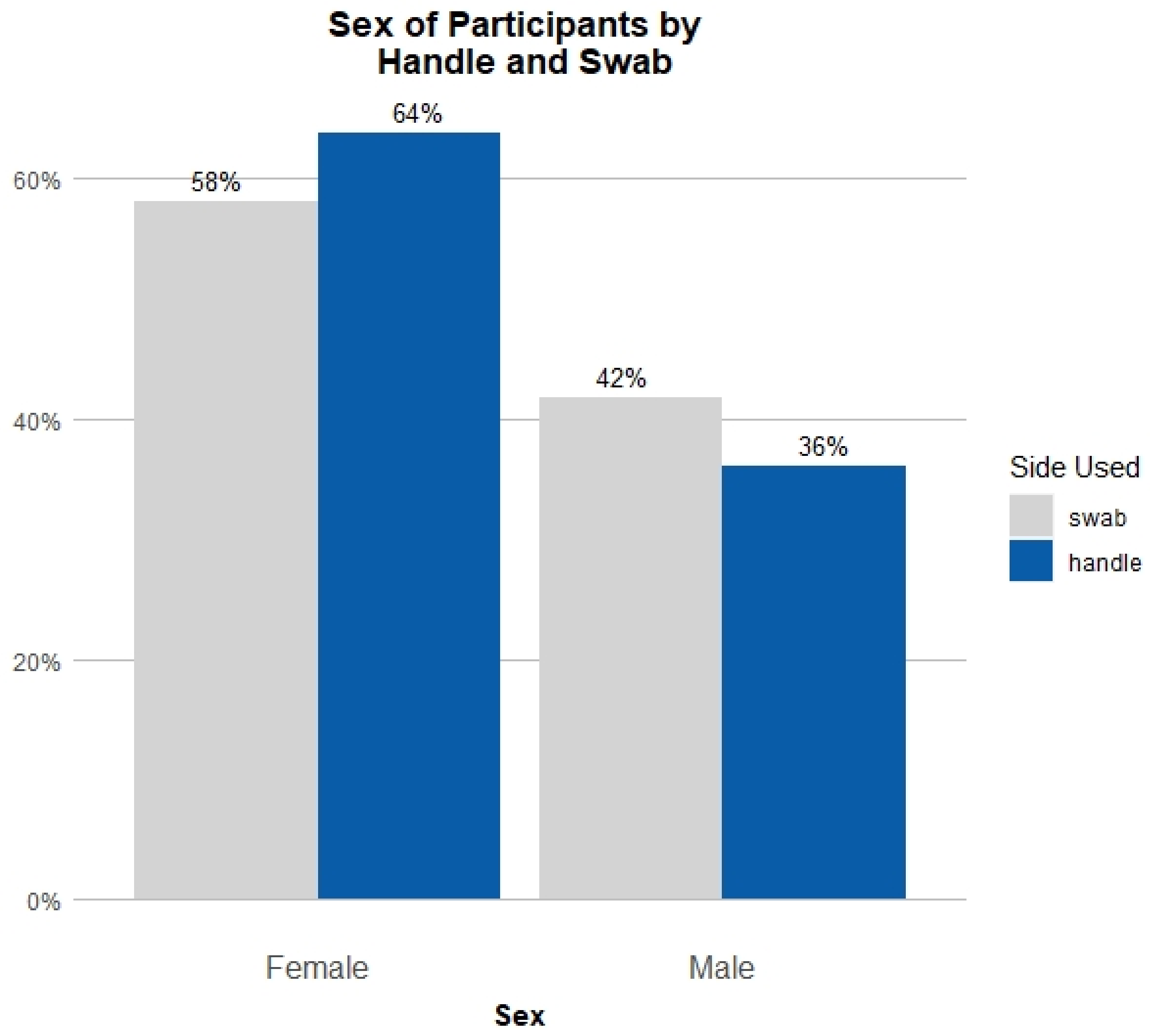


A chi-square test with Yates' continuity correction is used to compare rates of handle versus swab use between men and women.

```
# create a 2x2 dataframe in prep of chi-square test
dfSex <- data.frame(side_used = sex_group$side_used, sex = sex_group$sex_group)

# chi-square test
chisq.test(dfSex$sex,dfSex$side_used)

# display table
table(dfSex)
```

With a p-value of **0.22**, handle use did not differ significantly between female and male.

#### Data Analysis and Visualization for Income

Similarly to the last section, before a visualization is generated, participant incomes are labeled and samples lacking income data are removed.

```r
# new dataframe created
dfIncome <- participant_df
dfIncome <- dfIncome %>% filter (!is.na(income))
dfIncome %>% group_by(income) %>% summarise()

# income binnings are created
income_group <- dfIncome %>% mutate(income_group = case_when(
  income == "100k_125k" ~ '100-125k',
  income == "125k_150k" ~ '125-150k',
  income == "25k_50k" ~ '25-50k',
  income == "50k_75k" ~ '50-75k',
  income == "75k_100k" ~ '75-100k',
  income == "dont_know" ~ "NA",
  income == "dont_say" ~ "NA",
  income == "less_25k" ~ '<25k',
  income == "more_150k" ~ '>150k',))
```

Incomes are compared using a chi-squared test.

```r
chisq.test(dfIncome$income,dfIncome$side_used)
```

With a p-value of **0.095**, handle use did not differ significantly by income.

Prior to visualization, a dataframe called income_hist is prepared with frequency for each income group based on side used.

```r
# dataframe created with frequency and count
income_hist <- income_group %>%
  group_by(side_used,income_group,.drop=FALSE) %>%
  dplyr::summarise(n = n()) %>%
  complete(side_used, fill=list(count_a =0))%>%
  mutate(freq = n / sum(n))%>%
  ungroup()
sum(income_hist$freq)

# QC-ing of the dataframe (factor)
income_group %>% group_by(income_group) %>% summarise()
income_hist$income_group <- factor(income_hist$income_group, levels =
                                     c("<25k","25-50k","50-75k","75-100k",
                                       "100-125k","125-150k",">150k","NA"))
income_hist$side_used <- factor (income_hist$side_used, levels = c("swab","handle"))
```

With this dataframe, a bar plot can be generated.

```r
ggplot(income_hist, aes(x=income_group,y=freq,fill=side_used, width = 0.8))+
  geom_bar(stat="identity",position = position_dodge(preserve="single"))+
  scale_y_continuous(labels = scales::percent)+
  theme(panel.background = element_rect(fill = "white"),
        panel.grid.major.y = element_line(size = 0.5, linetype = 'solid', color = "grey"),
        panel.grid.major.x = element_blank(),
        legend.position = "right",
        plot.title = element_text(face = "bold", hjust = 0.5),
        axis.text.x = element_text(size = 10, angle = 35, vjust=1),
```

```
      axis.title.x = element_text(face = "bold",vjust= -1),
      axis.ticks = element_blank(),
      axis.title.y = element_blank())+
geom_text(aes(label=scales::percent(freq,accuracy =1),y=freq), vjust=-0.5, size=3.5,
          position = position_dodge(width=.8))+
ggtitle("Household Income of Participants by Handle and Swab")+
scale_fill_manual(values = c("swab" = "lightgrey",
                             "handle" = "#095DA8"))+
labs(x="Income", fill = "Side Used")
```

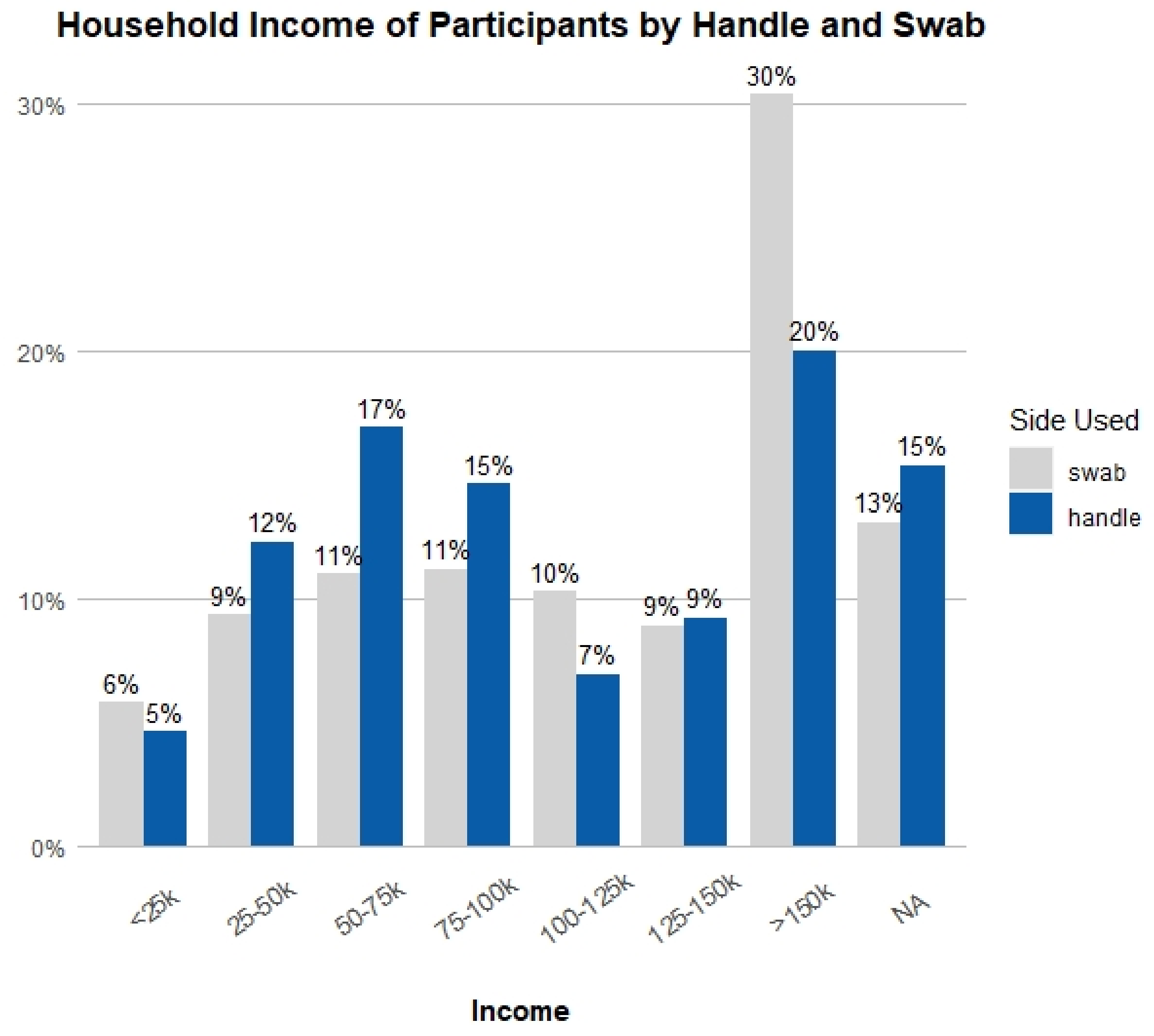


### SECTION 3: CALCULATING MEAN RNASEP CT VALUES

#### Creating the Dataframe

From the single dataset, we will create a new dplyr dataframe specifically for mean RNAse P Ct.

```r
crt_data <- SCAN_SFS_df
# create and modify dataframe to include only 4 columns
crt_data <- crt_data %>% subset(select = c("side_used","crt","sample_id","rack_id"))

# frop the NAs from the crt column
crt_data <- drop_na(crt_data, crt)

# convert to a data table
crt_data <- as.data.table(crt_data)
```

#### Reshaping Ct Data of Handle versus Swab

To perform a t-test, the data must be reshaped to separate handle Cts and swab Cts into separate columns.

```r
# new dataframe created where crt values are assigned, dependent on type
crt_cast <- dcast(crt_data, sample_id ~ side_used, value.var = "crt", fun.aggregate = sum)

# Convert null values coerced to 0 during casting back to null
crt_cast$handle <- na_if(crt_cast$handle,0)
crt_cast$swab <- na_if(crt_cast$swab,0)
```

#### Calculating Mean with 95% Confidence Intervals for Handle versus Swab

The mean and 95% confidence intervals are calculated for handle versus swab samples.

```r
# calculate mean RNAse P CRTs
handle_x <- mean(crt_cast$handle,na.rm = TRUE)
swab_x <- mean(crt_cast$swab, na.rm = TRUE)

# calculate 95% confidence intervals
handle_ci = qnorm(0.975)*sd(na.omit(crt_cast$handle)) /
  sqrt(length(na.omit(crt_cast$handle)))
swab_ci <- qnorm(0.975)*sd(na.omit(crt_cast$swab)) /
  sqrt(length(na.omit(crt_cast$swab)))

# to find the upper and lower ranges for the mean based on the 95% CI
handle_lower = handle_x - handle_ci
handle_upper = handle_x + handle_ci
swab_lower = swab_x - swab_ci
swab_upper = swab_x + swab_ci
```

The mean for handle is **18.2** with 95% CI of **0.76**.

The mean for swab is **16.3** with 95% CI of **0.049**.

#### Comparison of Ct Values

The RNAse P Ct values are compared between handle and swab using a two-tailed Welch's two-sample t-test.

```r
t.test(crt_cast$handle,crt_cast$swab)
```

With a p-value less than **0.01**, the mean Ct for samples taken with the handle was significantly higher than for samples taken with the swab.

#### Creating a Boxplot for Mean RNAseP Ct Values

RNAse P CRT values for handle versus swab are visualized using a boxplot also containing sample sizes for swabs versus handles.

```r
# calculate N's
crt_data %>% group_by(side_used) %>% summarise(n=n())

boxplot <- crt_data %>%
  ggplot(aes(x=side_used,y=crt,fill=side_used))+
  geom_boxplot()+
  scale_fill_manual(values = c("swab" = "lightgrey",
                               "handle" = "#095DA8"))+
  theme_light()+
  theme(panel.background = element_rect(fill = "white"),
        panel.grid.major.y = element_line(size = 0.5, linetype = 'solid', color = "grey"),
        legend.position = "none",
        plot.title = element_text(face = "bold", hjust = 0.5),
        axis.title.x = element_text(face = "bold"),
        axis.title.y = element_text(face = "bold"),
        axis.text = element_text(size = 11))+
  ggtitle("Human RNAse P CRT Values by Handle and Swab")+
  scale_y_continuous("CRT", breaks = c(10,15,20,25,30), labels = c("10","15","20","25","30"))+
  geom_hline(yintercept=28, linetype = "longdash",color="#4D4D4D")+
  #  annotate("rect", xmin= 1.26,xmax=1.74,ymin=27.5,ymax=28.5,fill="white")+
  #  annotate("text", y=28,x=1.5, label="Limit of Detection", color = "black")+
  annotate("text",x=1.3,y=21.7, label = "N=99")+
  annotate("text",x=2.27,y=18.8, label = "N=11904")+
  xlab("Side used")+
  ylab("CRT")
print(boxplot)
```

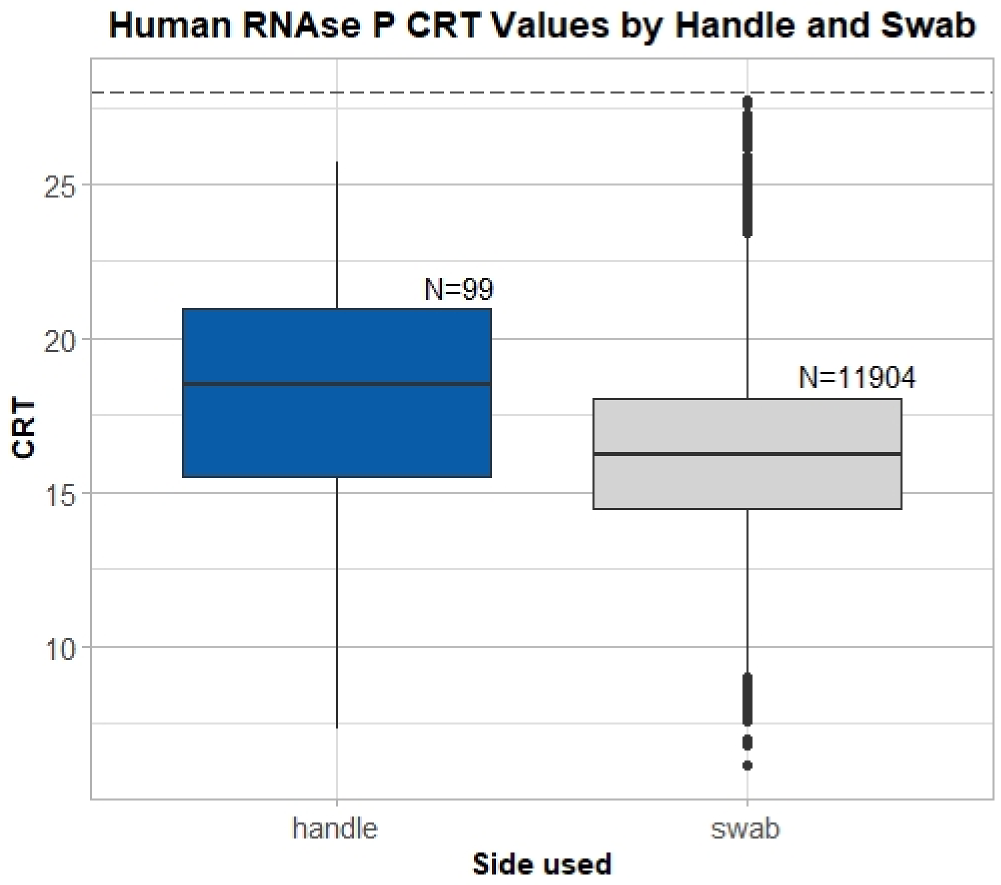


#### Creating a Dot Plot

To visualize how handle Ct values fall within the general range of swab Ct values, a dot plot is generated. Plates containing at least one handle sample are arranged by date and color coded by side used.

```
# rack id of the samples that was collected via handle
handle_racks <- data.table(rack_id = na.omit(unique(ifelse(crt_data$side_used == "handle",
                                                           crt_data$rack_id,NA))))

# rack id and crt of samples collected via swab
swabs_within_handle_racks <- left_join(handle_racks,crt_data,by="rack_id")
swabs_within_handle_racks <-
  swabs_within_handle_racks[grepl("swab", swabs_within_handle_racks$side_used),]

# rack id and crt of samples collected via handle
handles_within_handle_racks <- crt_data[grepl("handle", crt_data$side_used),]

# dot plot generation
p <- ggplot()+
  geom_point(
    data=swabs_within_handle_racks,
    aes(x=as.character(rack_id),y=crt,color="swab"),
    size=1)+
  geom_point(
```

```
    data=handles_within_handle_racks,
    aes(x=as.character(rack_id),y=crt,color="handle"),
    size=1)+
  theme_bw()+
  theme(panel.grid.minor.y = element_blank(),
        panel.border = element_rect(color="lightgrey"),
        plot.title = element_text(face = "bold", hjust = 0.5),
        axis.title.x = element_text(face = "bold"),
        axis.title.y = element_text(face = "bold"),
        axis.text.x = element_text(face = "bold",size = 7),
        axis.ticks = element_blank(),
        #         plot.margin = unit(c(0.2,1,0.2,0.2), "in"),
        #         legend.position = c(1.07,0.5),
        #         legend.background = element_rect(color = "lightgrey"),
        axis.line.x = element_blank())+
  ylim(10,30)+
  scale_x_discrete(breaks = c("021","054","100","133","206"), labels = c("Dec","Jan","Feb","Mar","Apr")
  ggtitle("Human RNAse P CRT Values by Handle and Swab")+
  #  geom_text(aes(x="21",y=9.5, label="Dec 2019"))+
  scale_color_manual(name = "Side Used", values = c("swab" = "lightgrey",
                                                  "handle" = "#095DA8"))+
  xlab("Date")+
  ylab("CRT")
print(p)
```

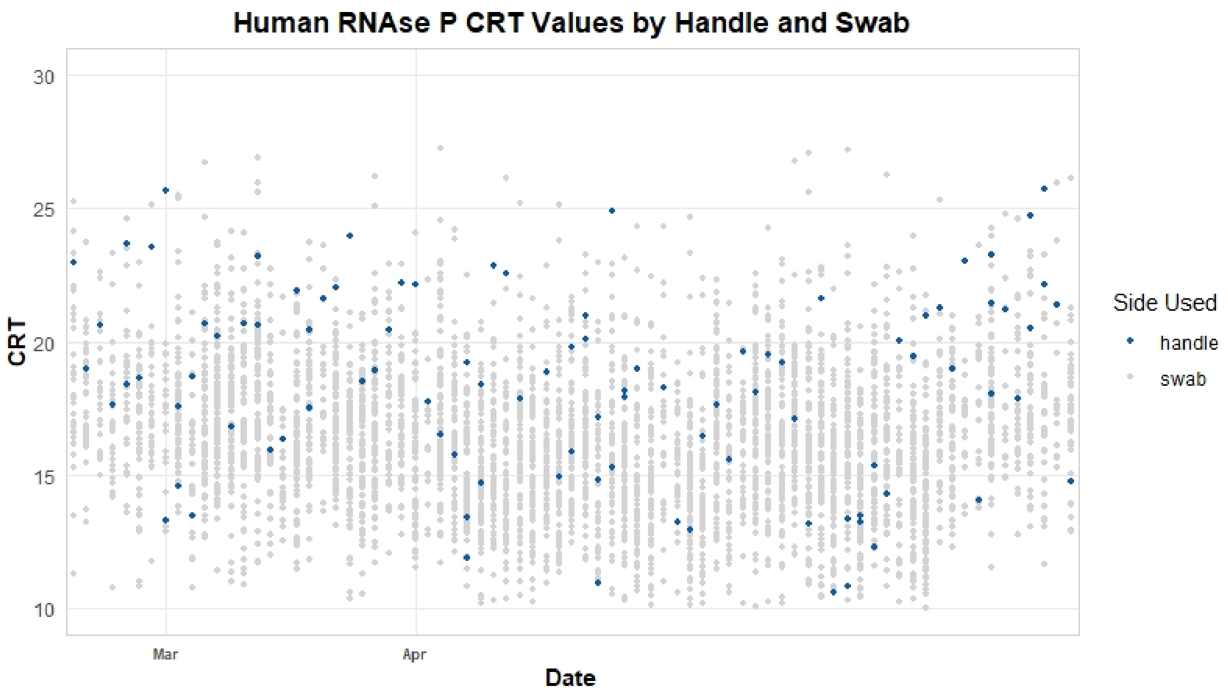

### SECTION 4: CALCULATING PATHOGEN DETECTION

#### Creating the Dataframe

From the single dataset, we will create a new dplyr dataframe specifically for pathogen detection, selecting samples for which RNAse P was detected.

```
# create new dataframe
path_detect_df <- SCAN_SFS_df

# select appropriate columns
path_detect_df <- select(path_detect_df, !c(rack_id, crt, sex, confidence, discomfort, age,
                                            income, aliquoted_date, side_used.y))

# filter and remove amples with Rnase P failures
ctrldet <- path_detect_df %>% filter(!is.na(rnasep_detected))
ctrldet <- ctrldet %>% filter(rnasep_detected == "Detected")
```

#### Creating the Table and Subset Dataframes

A table can be created with new columns that will display total counts of detected versus not detected by pathogen. The values in the initial table default to 0.

```
# get pathogen names from columns
detcol <- colnames(ctrldet[4:19])

# create dataframe with new column names, preset to value of 0
TFdetected <- data.frame("Pathogen" = detcol, "Handle_Present" = 0,
                         "Handle_Not_Present" = 0, "Swab_Present" = 0,
                         "Swab_Not_Present" = 0, stringsAsFactors = FALSE)
```

The ctrldet dataframe can be split into two separate dataframes, based on handle-type versus swab-type. As samples taken before 2020 were not tested for SARS-CoV-2 and many samples were tested solely on the LDT SARS-CoV-2 assay, the data for SARS-CoV-2 will have a different denominator.

```
# group samples by handles vs. swabs
# handle
ctrldetH <- ctrldet %>% filter(side_used=="handle")

# swab
ctrldetS <- ctrldet %>% filter(side_used=="swab")

# covid array
sars_set <- ctrldet %>% filter(!is.na(ctrldet$SARSCOV2))
## handle
sars_set_H <- sars_set %>% filter(side_used=="handle")
## swab
sars_set_S <- sars_set %>% filter(side_used=="swab")

# open array
oa_set <- ctrldet %>% filter(!is.na(ctrldet$Adenovirus))
## handle
```

```
oa_set_H <- oa_set %>% filter(side_used=="handle")
## swab
oa_set_S <- oa_set %>% filter(side_used=="swab")
```

The dataframes created above are used to fill in the TFdetected table for each pathogen and its rate of detection.

```
# fill table with numbers of detection for each pathogen for handles vs. swabs
for(row in 1:nrow(TFdetected)){
  TFdetected$Handle_Present[row] <-
    nrow(oa_set_H[oa_set_H[,TFdetected$Pathogen[row]]==TRUE,])
  TFdetected$Handle_Not_Present[row] <-
    nrow(oa_set_H[oa_set_H[,TFdetected$Pathogen[row]]==FALSE,])
  TFdetected$Swab_Present[row] <-
    nrow(oa_set_S[oa_set_S[,TFdetected$Pathogen[row]]==TRUE,])
  TFdetected$Swab_Not_Present[row] <-
    nrow(oa_set_S[oa_set_S[,TFdetected$Pathogen[row]]==FALSE,])
}

# adjust SARS-CoV-2 numbers using totals found in the sars_set, sars_set_H, and sars_set_S
TFdetected$Handle_Present[TFdetected$Pathogen=="SARSCOV2"] <-
  nrow(sars_set_H[sars_set_H$SARSCOV2==TRUE,])
TFdetected$Handle_Not_Present[TFdetected$Pathogen=="SARSCOV2"] <-
  nrow(sars_set_H[sars_set_H$SARSCOV2==FALSE,])
TFdetected$Swab_Present[TFdetected$Pathogen=="SARSCOV2"] <-
  nrow(sars_set_S[sars_set_S$SARSCOV2==TRUE,])
TFdetected$Swab_Not_Present[TFdetected$Pathogen=="SARSCOV2"] <-
  nrow(sars_set_S[sars_set_S$SARSCOV2==FALSE,])
```

The detection rate is inserted into the table, separated by two columns, handle versus swab. The SARS-COV detection rate is adjusted appropriately to reflect the different denominator.

```
# add decimal form of rates of detection for each pathogen for handle
TFdetected <- TFdetected %>% mutate(dec_rate_H = Handle_Present/nrow(oa_set_H))

# add decimal form of rates of detection for each pathogen for swab
TFdetected <- TFdetected %>% mutate(dec_rate_S = Swab_Present/nrow(oa_set_S))

# adjust SARS-CoV-2 rates based on totals found in s_set, sars_set_H, and sars_set_S
## handle
TFdetected$dec_rate_H[TFdetected$Pathogen=="SARSCOV2"] <-
  TFdetected$Handle_Present[TFdetected$Pathogen=="SARSCOV2"]/nrow(sars_set_H)
## swab
TFdetected$dec_rate_S[TFdetected$Pathogen=="SARSCOV2"] <-
  TFdetected$Swab_Present[TFdetected$Pathogen=="SARSCOV2"]/nrow(sars_set_S)
```

The dataframe now shows total counts of presence and absence for each pathogen, along with its rate of detection. The finalized table will be created in the next section.

#### Calculating P-Value

The rate of pathogen detection between handle and swab is compared using a paired t-test.

```
# run paired t-test on rates of detection
t.test(as.numeric(TFdetected$dec_rate_H),as.numeric(TFdetected$dec_rate_S),paired=TRUE)
```

With a p-value of **0.51**, the two sides did not differ in the rate at which they detected pathogens.

#### Creating Finalized Tables

The final table is prepared by first relabeling the pathogens with appropriate naming.

```
# rename pathogens and rate columns for new table
TFdetected2 <- TFdetected
TFdetected2$Pathogen[TFdetected2$Pathogen=="Flu_A"] <- "Influenza A"
TFdetected2$Pathogen[TFdetected2$Pathogen=="Flu_B"] <- "Influenza B"
TFdetected2$Pathogen[TFdetected2$Pathogen=="Flu_C"] <- "Influenza C"
TFdetected2$Pathogen[TFdetected2$Pathogen=="RSV"] <- "Respiratory Syncytial Virus"
TFdetected2$Pathogen[TFdetected2$Pathogen=="SARSCOV2"] <- "SARS-CoV-2*"
TFdetected2$Pathogen[TFdetected2$Pathogen=="Seasonal_Coronavirus"] <- "Seasonal Coronavirus"
TFdetected2$Pathogen[TFdetected2$Pathogen=="C.Pneumoniae"] <- "C. pneumoniae"
TFdetected2$Pathogen[TFdetected2$Pathogen=="S.Pneumoniae"] <- "S. pneumoniae"
TFdetected2$Pathogen[TFdetected2$Pathogen=="M.Pneumoniae"] <- "M. pneumoniae"
names(TFdetected2)[names(TFdetected2) == "dec_rate_H"] <- "pct_rate_H"
names(TFdetected2)[names(TFdetected2) == "dec_rate_S"] <- "pct_rate_S"
```

Next, the table should show total detected counts along with percent detected per pathogen. A percent function is used, and SARS-CoV-2 rates are updated from the earlier dataframes.

```
# create percent function
percent <- function(x, digits = 1, format = "f", ...) {
  paste0(formatC(100 * x, format = format, digits = digits, ...), "%")
}


# fill in percentages on TFdetected2 table
for (row in 1:nrow(TFdetected)){
  TFdetected2$pct_rate_H[row] <-
    percent(TFdetected2$Handle_Present[row]/nrow(oa_set_H))
  TFdetected2$pct_rate_S[row] <-
    percent(TFdetected2$Swab_Present[row]/nrow(oa_set_S))
}


# recalculate SARS-CoV-2 rates based on totals from s_set, sars_set_H, and sars_set_S
## handle
TFdetected2$pct_rate_H[TFdetected2$Pathogen=="SARS-CoV-2*"] <-
  percent(TFdetected2$Handle_Present[TFdetected2$Pathogen=="SARS-CoV-2*"]/nrow(sars_set_H))
## swab
TFdetected2$pct_rate_S[TFdetected2$Pathogen=="SARS-CoV-2*"] <-
  percent(TFdetected2$Swab_Present[TFdetected2$Pathogen=="SARS-CoV-2*"]/nrow(sars_set_S))
```

With the raw table prepared, the final table with proper formatting is generated. This table sorts the pathogen alphabetically while and moves bacterial pathogens to the bottom of the table. The table is then saved as a csv as pathogen_detection_published_table.

```
# get pathogen names from TFdetected2
detcol2 <- TFdetected2$Pathogen

# create finalized table for publication
pcttable <- data.frame("Pathogen"= detcol2)


# fill in values from detection rate table and format for publication
pcttable <- pcttable %>% mutate(Handle_Present =
                                  paste(paste(TFdetected2$Handle_Present,
                                              TFdetected2$pct_rate_H,
                                              sep=" ("), "",sep=")"))
pcttable <- pcttable %>% mutate(Swab_Present =
                                  paste(paste(TFdetected2$Swab_Present,
                                              TFdetected2$pct_rate_S,
                                              sep=" ("), "",sep=")"))


# sort table by pathogen (alphabetically) and move bacteria to the bottom
pcttable <- pcttable[order(pcttable$Pathogen),]
table_top <- pcttable[-grep("pneumoniae",pcttable$Pathogen),]
table_bottom <- pcttable[grep("pneumoniae",pcttable$Pathogen),]
final_pcttable <- rbind(table_top, table_bottom)


# display table and write as csv
view(final_pcttable)
write.csv(pcttable3, 'pathogen_detection_published_table.csv', row.names = FALSE)
```
